# Low-Dose Tenecteplase for Acute Ischemic Stroke Treatment in Aging Patients DATE-AGING Trial: Rationale and Design

**DOI:** 10.64898/2026.10.08.26361416

**Authors:** Xianhua Hou, Shuai Mi, Xi Chen, Yuxuan He, Lin Chen, Qu Liu, Junhua Wu, Tao Lv, Liangbo Kong, Ruchuang Xu, Min Wu, Duolao Wang, Zhenhua Zhou, DATE-AGING investigators

## Abstract

**Background:** Despite the high stroke burden in the elderly, their thrombolysis utilization is suboptimal partly due to heightened hemorrhagic risk. Observational data suggest that low-dose thrombolysis may offer comparable efficacy to standard-dose with a trend toward reduced symptomatic intracranial hemorrhage (sICH), yet high-level evidence for low-dose thrombolysis in this broad population remains absent.

**Objective:** To evaluate the safety and efficacy of low-dose intravenous tenecteplase thrombolysis in elderly stroke patients within 4.5 hours of ischaemic stroke onset.

**Methods and design:** The DATE-AGING trial is a multicenter, prospective, randomized, open-label, blinded-end point (PROBE) non-inferiority study. Eligible participants are elderly patients (aged ≥70 years) with acute ischemic stroke who meet current guideline recommendations for intravenous thrombolysis within 4.5 hours of symptom onset will be randomly assigned to Low-dose Tenecteplase group (0.175 mg/kg, max 17.5 mg) or Standard-dose Tenecteplase group (0.25 mg/kg, max 25 mg). The primary outcome is excellent functional outcome, defined as modified Rankin Scale(mRS) 0-1 at 90 days. Safety outcomes include sICH within 36 hours and mortality at 90 days.

**Discussions:** The DATE-AGING trial is intended to provide high-quality evidence for low-dose tenecteplase intravenous thrombolysis in elderly patients with acute ischemic stroke.

**Trial registry number:** NCT07294209.

## Introduction

Acute ischemic stroke (AIS) is one of the most common diseases with significant morbidity, mortality, and disability^1^. With the accelerating aging of China’s population, the age distribution of stroke patients has continued to shift upward, and the incidence of stroke among individuals over 70 years of age has increased rapidly^2^.

Multiple studies have identified early intravenous thrombolysis as the most effective clinical treatment for elderly patients with AIS^3–5^. Subgroup analyses from the Third International Stroke Trial (IST-3) demonstrated that thrombolysis administered within 4.5 hours of symptom onset improved neurological outcomes in elderly patients^6^. Another systematic review reported that intravenous alteplase administered within 4.5 hours of symptom onset was beneficial even for patients aged over 80 years^3^. Based on these results, current guidelines from the American Heart Association/American Stroke Association (AHA/ASA), the Chinese Stroke Association, and the European Stroke Organisation (ESO) all support the use of intravenous alteplase within the 4.5 hour time window for acute ischemic stroke patients over 80 years of age.^7–9^. Despite the proven efficacy, intravenous thrombolysis is substantially underutilized in the elderly stroke population. In a large-scale nationwide registry study from China, Li et al^10^ reported that patients aged ≥80 years who were eligible for thrombolysis, only 28.1% ultimately received alteplase. Notably, alteplase use was associated with a more than threefold increase in the risk of sICH. Alshekhlee et al^11^ analyzed a large US national database and found that elderly patients with acute ischemic stroke had a significantly higher risk of intracranial hemorrhage after thrombolysis, yet they were less likely to receive intravenous thrombolysis. Menon et al^12^ quantified the impact of age on post-thrombolysis bleeding risk, demonstrating that each 10-year increase in age was associated with 26% higher odds of sICH in their multivariate regression model. This pronounced safety concern likely exacerbates clinical hesitation, which may in turn account for the observation that thrombolysis utilization rates are substantially lower in elderly patients compared with the younger counterparts. Tenecteplase, a genetically modified variant of tissue plasminogen activator, offers enhanced fibrin specificity and a prolonged half-life, enabling single-bolus administration that simplifies thrombolytic delivery and reduces door-to-needle time in AIS. In the TRACE-2 post-hoc analysis, tenecteplase (0.25mg/kg) showed comparable functional outcomes to alteplase but did not reduce sICH (4.0% vs. 3.3%)^13^. This unresolved safety concern limits its broader adoption in this vulnerable population.

In recent years, the low-dose (0.6 mg/kg) alteplase regimen has been increasingly explored as an alternative to standard-dose thrombolysis particular in Asian clinical practice^14–16^. ENCHANTED trial^17^, a large randomized controlled trial to compare low-dose (0.6 mg/kg) versus standard-dose (0.9 mg/kg) intravenous alteplase. The study found that low-dose alteplase significantly reduced the risk of sICH but did not meet the non-inferiority criterion for the primary outcome of death or disability at 90 days. In a meta-analysis of 12 studies comprising 7,686 patients, Cheng et al.^18^ found that low-dose alteplase was comparable to standard-dose in improving functional outcomes and reducing mortality while significantly reducing the incidence of sICH. Regarding low-dose thrombolysis in elderly patients, Chao et al.^19^ reported that lower-dose therapy was associated with significantly lower sICH and mortality rates than standard-dose treatment in Chinese stroke patients aged ≥70 years alongside improved functional independence. These findings support the potential of dose reduction as a strategy to improve the risk-benefit balance in this vulnerable population. The risk-benefit paradox-where elderly stroke patients face high bleeding risk, yet low-dose thrombolysis may reduce sICH has driven off-label use of reduced-dose alteplase^20^. These prescribing practices represent clinical efforts to balance efficacy and safety in elder patients. However, whether dose-reduced thrombolysis offers a more favorable risk-benefit profile than the standard-dose regimen in elderly stroke patients remains unclear. Given the molecular pharmacological advantages of tenecteplase, low-dose tenecteplase may offer a favorable efficacy–safety balance. Thus, DATE-AGING trial is designed to explore the efficacy and safety of low-dose tenecteplase thrombolysis in elderly patients with acute ischemic stroke.

## Methods

### Study design

The DATE-AGING trial is a multicenter, prospective, randomized, open-label, blinded-endpoint (PROBE) non-inferiority study (Figure 1). It is an academic trial designed by the principal investigators and a steering committee consisting of experts in cerebrovascular diseases. This trial adheres to the Declaration of Helsinki and has been approved by the ethics committee of Chongqing Southwest Hospital, Army Medical University and all participating centers. Written informed consent will be obtained from all patients or their legal representatives after they receive detailed information about the purpose of the trial, possible benefits and risks, and the rights/obligations. This study includes approximately 50 stroke centers in China. These sites are able to offer round-the-clock comprehensive stroke care to patients with AIS. This trial has been registered at https://clinicaltrials.gov (NCT07294209). The trial flowchart is shown in Figure 2.

**Figure 1.**
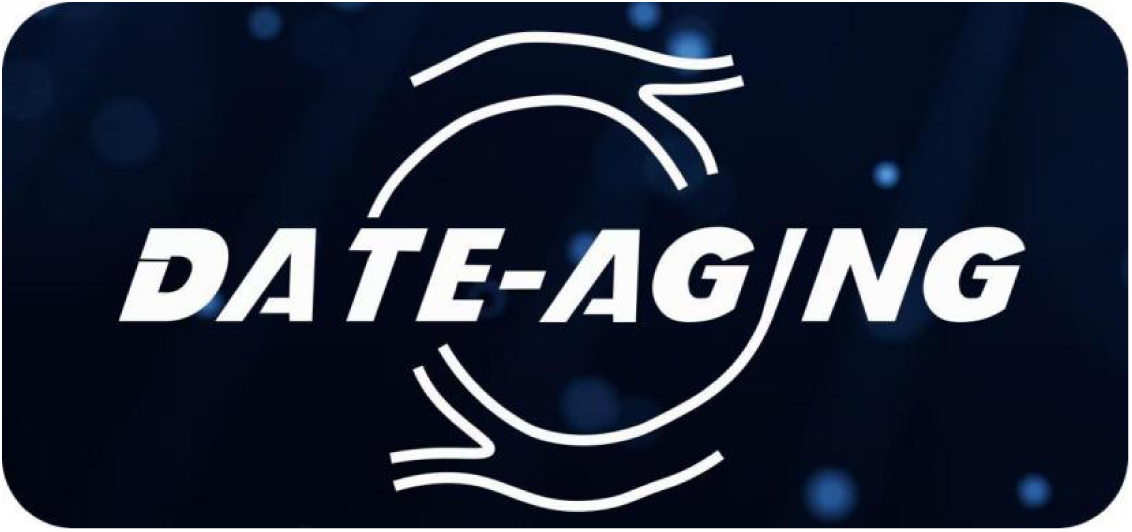
DATE-AGING trial logo.

**Figure 2.**
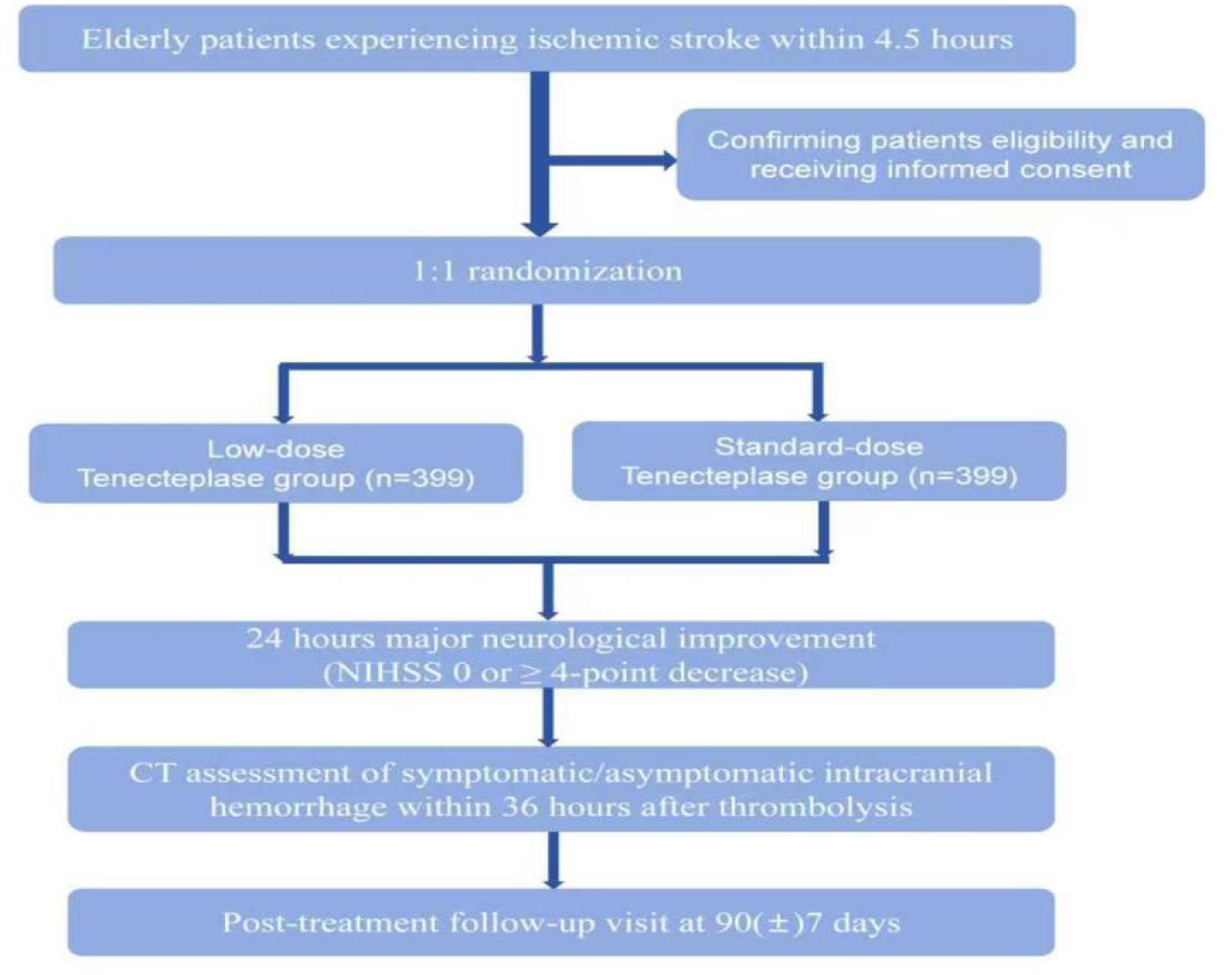
Flow chart of the DATE-AGING trial.

## Participant population

### Inclusion Criteria

1. Age≥70 years;
2. Diagnosis of ischaemic stroke with a measurable neurological deficit on National Institutes of Health Stroke Scale (NIHSS) (1≤NIHSS≤25); if NIHSS<4, patients have to be with at least a measurable deficit on motor power (upper or lower limbs≥1);
3. Stroke symptoms should have been present for at least 30 minutes (min) without significant improvement prior to randomisation;
4. Thrombolytic therapy can be initiated within 4.5 Hour(s) (h) of Acute ischaemic stroke (AIS) onset;
5. Patients with premorbid modified Rankin Scale (mRS) 0 or 1;
6. Informed consent from the patient or surrogate.

### Exclusion Criteria

1. Imaging demonstrates multi-lobar infarction (hypodensity>1/3 cerebral hemisphere);
2. Acute bleeding diathesis or allergy to tenecteplase, including but not limited to:
  ⮚ Known genetic predisposition to bleeding or significant bleeding disorder at present or within the past 6 months
  ⮚ Administration of heparin within the previous 48h and activated partial thromboplastin time (aPTT) exceeding the upper limit of normal for laboratory measurement
  ⮚ Current use of vitamin K based oral anticoagulants (e.g. warfarin) and a prolonged prothrombin time (International normalised ratio (INR) > 1.7 or Prothrombin time (PT)>15 seconds (s)) or current use of novel oral anticoagulants (i.e. dabigitran, rivaroxiban, or apixiban) with prolongation of activated partial thromboplastin time (aPTT) and/or PT above the upper limit of the local laboratory reference range
  ⮚ Platelet count of below 100×10^9^/ L
  ⮚ Any history of central nervous system damage (i.e. neoplasm, aneurysm, intracranial or spinal surgery)
  ⮚ Recent traumatic external heart massage or recent puncture of a non-compressive blood-vessel (e.g. subclavian or jugular vein puncture) within the past 10 days
  ⮚ Known history of suspected intracranial haemorrhage or suspected subarachnoid haemorrhage from aneurysm
  ⮚ Neoplasm with increased haemorrhagic risk
  ⮚ Documented ulcerative gastrointestinal disease during the last 3 months, oesophageal varices, arterial aneurysm, or arterial/venous malformations
  ⮚ History of significant trauma or major surgery within the past 3 months
  ⮚ Any known disorder associated with a significant increased risk of bleeding
3. Intracranial hemorrhage (including parenchymal hemorrhage, intraventricular hemorrhage, subarachnoid hemorrhage, subdural/extradural hematoma, etc.);
4. Blood glucose <2.8 mmol/L or >22.22 mmol/L;
5. After active antihypertensive treatment, hypertension is still not under control: systolic blood pressure≥180 mmHg, or diastolic blood pressure ≥100 mmHg;
6. Seizure at stroke onset;
7. Concurrent malignancy or severe systemic disease with an anticipated survival of less than 90 days;
8. Participation in other clinical trials within 3 months prior to screening;
9. Unsuitability or participation in this study as judged by the investigator may result in subjects being exposed to greater risk.

### Baseline measures

Patients with suspected AIS who present within 4.5 hours of symptom onset undergo evaluation for intravenous thrombolysis following neuroimaging. Demographic data, medical history, concomitant medications, and baseline laboratory parameters are recorded. Baseline neurological severity (NIHSS) and pre-stroke disability (mRS) are assessed by a trained and certified physician following a standardized rapid workflow. Patients scheduled for endovascular thrombectomy are eligible for enrollment in this study.

### Randomization and blinding

Eligible patients are randomly assigned in a 1:1 ratio to receive either low-dose or standard-dose intravenous tenecteplase. Randomization is performed centrally via a web-based, real-time interactive response technology system(lcsjcj.xnyy.cn) and is stratified by baseline NIHSS score (<6, 6–15, >15), with a randomization sequence generated using a validated pseudorandom number generator. The intravenous thrombolytic treatment is open-label. Investigators involved in subsequent clinical and imaging outcome assessments are blinded to treatment allocation.

## Treatment intervention

### Low-dose Tenecteplase group

Tenecteplase (0.175 mg/kg) is administered as a single intravenous bolus (over 5–10 seconds) immediately upon randomization, with a maximum dose of 17.5 mg.

### Standard-dose Tenecteplase group

Tenecteplase (0.25 mg/kg) is administered as a single intravenous bolus (over 5–10 seconds) immediately upon randomization, with a maximum dose of 25 mg.

All enrolled patients are monitored in the acute stroke unit and may be admitted to the intensive care unit if clinically indicated. All patients receive standardized medical treatment management and subsequent secondary preventive medications in accordance with the 2023 Chinese Guidelines for Diagnosis and Treatment of Acute Ischemic Stroke^8^.

### Primary outcome

Percentage of participants with mRS score of 0 or 1 at day 90(±7).

### Secondary outcomes Efficacy outcomes

1. Percentage of participants with mRS score of 0-2 at day 90(±7);
2. Distribution of mRS at day 90(±7);
3. Percentage of participants with major neurological improvement (NIHSS score of 0 or improvement of at least 4 points compared with baseline) at 24 hours;
4. EQ-5D-5L score at day 90(±7);
5. Percentage of participants with Barthel Index (BI) Score ≥95 at day 90(±7).

### Safety outcomes

1. Percentage of participants with symptomatic intracerebral haemorrhage (sICH) within 36 hours as defined by European Cooperative Acute Stroke Study (ECASS III);
2. Percentage of participants with mRS score of 5 or 6 at day 90(±7);
3. All-cause mortality within 90 days.

### Data safety monitoring board

An independent Data and Safety Monitoring Board (DSMB) will be established, comprising three experts-a neurologist, a neuroradiologist, and a biostatistician independent of the overall study statistician. The DSMB members will not be participants in the trial or have any affiliation with the study sponsors. The DSMB will convene regularly to oversee the progress of the trial. Furthermore, the DSMB will review the occurrence of serious adverse events to guarantee the safety of the patients.

### Clinical events committee

The Clinical events committee(CEC) consists of experienced neurologists who are not involved in the execution of the study. Membership of the CEC (including qualifications, roles and responsibilities) will be confirmed by the steering committee. All clinical end points are first determined by the investigator, then verified and adjudicated by the CEC based on clinical symptoms, laboratory tests and imaging data.

### Sample size

The noninferiority margin was prespecified at −4.5%, consistent with the −5% margin adopted in previous noninferiority trials of tenecteplase versus alteplase^21,22^. This non-inferiority margin is also less than the lower 95% CI bound of approximately 6% on the point estimate of alteplase versus control (placebo) effect size in data from Emberson and colleagues^23^. For sample size calculation, we referred to a retrospective cohort study comparing low-dose versus standard-dose thrombolysis in stroke patients aged ≥70 years (27.1% vs. 22.2% for mRS 0–1 at 90 days) ^24^. Assuming 85% power and a one-sided α of 0.025, the estimated sample size was 379 patients per group. After accounting for a 5% withdrawal or loss to follow-up, the final target enrollment was set at 798 patients.

## Statistical analyses

### Analysis populations

Efficacy analyses will primarily use the intention-to-treat (ITT) population. The per-protocol population will support secondary analyses. Safety analyses will use the safety population.

### Primary efficacy endpoint

The primary outcome is the proportion of patients with mRS 0-1 at 90 days. The treatment effect is the risk difference (experimental minus control). The primary analysis uses a generalized linear model (GLM) with binomial distribution and identity link to calculate risk difference and its one-sided 97.5% confidence interval (CI) or equivalently two-sided 95% CI. Non-inferiority will be claimed if the lower bound of the two sided 95% CI for the risk difference exceeds -4.5%. Adjusted risk difference for baseline NIHSS score, age, and time to drug administration will also be estimated using GLM. If the adjusted model does not converge, inverse probability of treatment weighting approach will be used. Missing primary outcome data will be handled by multiple imputation as the primary analysis approach, supported by various sensitivity analyses including worse-case scenario, best-case scenario, worst-best scenario, center-adjusted, and tipping-point analyses. Subgroup analyses will also be performed on pre-specified subgroup variables including age, baseline NIHSS, time to treatment, sex, atrial fibrillation, diabetes, use of antiplatelet agent and thrombectomy. All sensitivity analyses and subgroup analyses are exploratory.

### Secondary efficacy endpoints

Binary secondary outcomes will be analyzed using modified Poisson regression (with a log-Poisson generalized linear model) to calculate risk ratio as measurement of treatment effect. The ordinal mRS will be analyzed using proportional odds logistic regression to estimate the common odds ratio as a measure of treatment effect; the proportional odds assumption will be formally tested. If the proportional odds assumption is violated, generalized odds ratio will be calculated. EQ-5D-5L scores will be analyzed using linear regression to estimate the mean difference as a measure of treatment effect, with the win ratio^25^ serving as a backup approach if the normality assumption is violated. Both unadjusted and adjusted estimates (mean differences or win ratios) will be calculated. All secondary analyses will be performed on both the ITT population and the per-protocol (PP) population^25^. These analyses are exploratory in nature.

### Safety endpoints

Safety analyses will be conducted on the safety population. Mortality at 90 days will be analyzed using modified Poisson regression for risk ratios and Cox regression for hazard ratios. Symptomatic intracranial haemorrhage and mRS 5-6 at 90 days will be analyzed using modified Poisson regression. Adverse events will be summarized descriptively. No multiplicity adjustment is planned for safety.

### General methods

All analyses will be performed using SAS 9.4 or R 4.3.3 or higher. Continuous variables will be summarized by mean, SD, median, quartiles, and range; categorical variables by frequencies and percentages. No adjustment for multiple comparisons is applied to secondary or safety analyses. The complete statistical analysis plan, including all imputation algorithms, convergence strategies, and sensitivity analyses, will be described in detail and finalized before the database lockup. The trial results will be reported according to the CONSORT guidelines.

## Discussion

The DATE-AGING trial is a multicenter, prospective, randomized, open-label, blinded-end point (PROBE) non-inferiority study to evaluate the efficacy and safety of low-dose tenecteplase in elderly patients with acute ischaemic stroke. China’s rapidly ageing population is expected to drive a continued rise in the incidence of acute ischaemic stroke among elderly patients^2^. Although the AHA/ASA/ESO and Chinese guidelines currently recommend intravenous alteplase within 4.5 hours of symptom onset for AIS patients aged >80 years^7–9^, nearly two-thirds of eligible elderly patients still do not receive thrombolysis in clinical practice^10^. A major concern underlying this treatment gap is the substantially elevated risk of sICH after thrombolysis^12^. Advanced age is recognized as one of the important risk factors for hemorrhagic transformation after thrombolysis^11^, as it systematically amplifies bleeding risk through multiple interconnected mechanisms. First, aging increases blood-brain barrier permeability and microvascular fragility due to degeneration of endothelial cells and basement membranes, allowing easier extravasation of blood components after reperfusion^26^. Second, elderly patients accumulate a higher burden of cerebral small vessel disease, including white matter hyperintensities and cerebral microbleeds, which serve as structural vulnerabilities for subsequent hemorrhage^27^. Third, age often coexists with multiple comorbidities such as hypertension, atrial fibrillation, and renal impairment, as well as prior use of antiplatelet or anticoagulant medications, further elevating baseline bleeding risk^28^. Therefore, advanced age serves not merely as a chronological threshold but as a composite marker of accumulated biological vulnerability that shifts the risk-benefit balance of thrombolytic therapy.

Low-dose thrombolysis for acute ischemic stroke refers to the use of a reduced dose of a thrombolytic agent (e.g., 0.6 mg/kg alteplase instead of the standard 0.9 mg/kg) to achieve a better safety profile-particularly a lower risk of sICH-while preserving acceptable recanalization efficacy. The ENCHANTED trial ^17^showed that low-dose alteplase significantly reduced sICH compared with standard-dose alteplase, although it did not meet non-inferiority for the primary outcome of functional recovery. With regard to low-dose thrombolysis in elderly stroke patients, the TTT-AIS II study demonstrated that a lower dose of 0.6 mg/kg alteplase offers significant advantages for elderly East Asian patients compared with the standard 0.9 mg/kg dose. This lower-dose regimen was associated with significantly lower rates of sICH, reduced mortality, and better functional independence.^19^ These findings, despite inherent limitations of observational data, support low-dose therapy as a reasonable option in elderly stroke patients. This risk-benefit consideration-heightened bleeding susceptibility versus the potential safety gain of dose reduction-has fuelled off-label use of reduced-dose thrombolysis in clinical practice^20^. However, robust evidence from randomized controlled trials remains lacking, underscoring the urgent need for a dedicated study to clarify the safety and efficacy of this strategy specifically in the elderly population.

Tenecteplase is a recombinant tissue plasminogen activator with three amino-acid modifications (T103N, N117Q, and KHRR 296–299 AAAA) that confer higher fibrin specificity and a longer half-life^29^. These properties permit single-bolus administration. Although this bolus regimen offers logistical advantages and eliminates infusion-related errors, it also removes the opportunity to stop treatment midway should an adverse event occur, making accurate dose selection particularly critical. Previous dose-ranging trials of tenecteplase (0.1, 0.25, and 0.4 mg/kg) have identified 0.25 mg/kg as the dose with the most favourable efficacy–safety profile, a finding further supported by a recent meta-analysis showing superiority over alteplase for mRS 0-1, whereas the 0.1 and 0.4 mg/kg doses did not show significant benefit^30^. This evidence base has led to guideline endorsement of 0.25 mg/kg tenecteplase for intravenous thrombolysis^8^. Nevertheless, it is important to note that most of these trials did not specifically target elderly patients, who are known to have an elevated risk of haemorrhagic complications. Notably, the TRACE-2 post-hoc analysis reported that tenecteplase (0.25 mg/kg) yielded functional outcomes comparable to alteplase, yet failed to reduce sICH (4.0% vs. 3.3%)^13^. Therefore, low-dose tenecteplase (0.1-0.25 mg/kg) remains largely investigational in elderly patients, and the optimal dose for this population has yet to be firmly established. The treatment intervention for the present study was informed by both precedent and pharmacodynamic rationale. The low-dose tenecteplase regimen (0.175 mg/kg) was set at 70% of the standard dose (0.25 mg/kg)-a proportional reduction comparable to the two-thirds dose reduction used for low-dose alteplase in the ENCHANTED^17^ and TTT-AIS II^19^ studies. This empirical precedent is further reinforced by the pharmacodynamic advantages of tenecteplase over alteplase, including approximately 15-fold higher fibrin specificity and a six-fold longer half-life, theoretically supporting sustained thrombolytic activity despite dose reduction. Collectively, these findings provide a rationale for evaluating low-dose tenecteplase in the elderly population.

In conclusion, the DATE-AGING study will evaluate the efficacy and safety of low-dose tenecteplase in elderly stroke patients through a prospective, multicenter, randomized controlled trial. The findings from this study will help elucidate the feasibility of low-dose intravenous tenecteplase thrombolysis strategies in this high-risk sICH elderly population.

## Data Availability

All data produced in the present study are available upon reasonable request to the authors

## Acknowledgement

None

## References

1. Global, regional, and national burden of stroke and its risk factors, 1990-2021: a systematic analysis for the Global Burden of Disease Study 2021. The Lancet Neurology. 2024;23(10):973–1003.

2. Zhang X, Dai J, Li W, Yang Y. High-risk population and factors of stroke has changed among middle-aged and elderly Chinese-Evidence from 1989 to 2015. Front Public Health. 2023;11:1090298.

3. Bluhmki E, Danays T, Biegert G, Hacke W, Lees KR. Alteplase for Acute Ischemic Stroke in Patients Aged >80 Years: Pooled Analyses of Individual Patient Data. Stroke. 2020;51(8):2322–2331.

4. Reuter B, Gumbinger C, Sauer T, et al. Intravenous thrombolysis for acute ischaemic stroke in the elderly: data from the Baden-Wuerttemberg stroke registry. Eur J Neurol. 2016;23(1):13–20.

5. Emberson J, Lees KR, Lyden P, et al. Effect of treatment delay, age, and stroke severity on the effects of intravenous thrombolysis with alteplase for acute ischaemic stroke: a meta-analysis of individual patient data from randomised trials. Lancet. 2014;384(9958):1929–1935.

6. group ISTc, Sandercock P, Wardlaw JM, et al. The benefits and harms of intravenous thrombolysis with recombinant tissue plasminogen activator within 6 h of acute ischaemic stroke (the third international stroke trial [IST-3]): a randomised controlled trial. Lancet. 2012;379(9834):2352–2363.

7. Powers WJ, Rabinstein AA, Ackerson T, et al. Guidelines for the Early Management of Patients With Acute Ischemic Stroke: 2019 Update to the 2018 Guidelines for the Early Management of Acute Ischemic Stroke: A Guideline for Healthcare Professionals From the American Heart Association/American Stroke Association. Stroke. 2019;50(12):e344–e418.

8. Liu L, Li Z, Zhou H, et al. Chinese Stroke Association guidelines for clinical management of ischaemic cerebrovascular diseases: executive summary and 2023 update. Stroke Vasc Neurol. 2023;8(6):e3.

9. Berge E, Whiteley W, Audebert H, et al. European Stroke Organisation (ESO) guidelines on intravenous thrombolysis for acute ischaemic stroke. European stroke journal. 2021;6(1):I–lxii.

10. Li C, Jiang Y, Gu HQ, et al. Characteristics, temporal trends and outcomes of intravenous thrombolysis in Chinese patients aged>80 years who had a stroke. Stroke Vasc Neurol. 2025;10(4):431–440.

11. Alshekhlee A, Mohammadi A, Mehta S, et al. Is thrombolysis safe in the elderly?: analysis of a national database. Stroke. 2010;41(10):2259–2264.

12. Menon BK, Saver JL, Prabhakaran S, et al. Risk score for intracranial hemorrhage in patients with acute ischemic stroke treated with intravenous tissue-type plasminogen activator. Stroke. 2012;43(9):2293–2299.

13. Xiong Y, Wang L, Pan Y, et al. Tenecteplase versus alteplase for acute ischaemic stroke in the elderly patients: a post hoc analysis of the TRACE-2 trial. Stroke Vasc Neurol. 2025;10(1):112–119.

14. Chen CH, Hsieh CY, Lai TB, Chuang MT, Chen WL, Sun MC. Optimal dose for stroke thrombolysis in Asians: low dose may have similar safety and efficacy as standard dose. Journal of thrombosis and haemostasis : JTH. 2012;10(7):1270–1275.

15. Miyamoto S, Ogasawara K, Kuroda S, et al. Japan Stroke Society Guideline 2021 for the Treatment of Stroke. International journal of stroke : official journal of the International Stroke Society. 2022;17(9):1039–1049.

16. Wang X, Li J, Moullaali TJ, et al. Low-dose versus standard-dose alteplase in acute ischemic stroke in Asian stroke registries: an individual patient data pooling study. International journal of stroke : official journal of the International Stroke Society. 2019;14(7):670–677.

17. Anderson CS, Robinson T, Lindley RI, et al. Low-Dose versus Standard-Dose Intravenous Alteplase in Acute Ischemic Stroke. N Engl J Med. 2016;374(24):2313–2323.

18. Cheng JW, Zhang XJ, Cheng LS, et al. Low-Dose Tissue Plasminogen Activator in Acute Ischemic Stroke: A Systematic Review and Meta-Analysis. Journal of stroke and cerebrovascular diseases : the official journal of National Stroke Association. 2018;27(2):381–390.

19. Chao AC, Liu CK, Chen CH, et al. Different doses of recombinant tissue-type plasminogen activator for acute stroke in Chinese patients. Stroke. 2014;45(8):2359–2365.

20. Mao XL, He SS, Zhang YX, et al. Low-dose vs. standard-dose alteplase for acute ischemic stroke: a real-world single-center retrospective study. Frontiers in neurology. 2025;16:1651307.

21. Menon BK, Buck BH, Singh N, et al. Intravenous tenecteplase compared with alteplase for acute ischaemic stroke in Canada (AcT): a pragmatic, multicentre, open-label, registry-linked, randomised, controlled, non-inferiority trial. Lancet. 2022;400(10347):161–169.

22. Muir KW, Ford GA, Ford I, et al. Tenecteplase versus alteplase for acute stroke within 4·5 h of onset (ATTEST-2): a randomised, parallel group, open-label trial. The Lancet Neurology. 2024;23(11):1087–1096.

23. Emberson J, Lees KR, Lyden P, et al. Effect of treatment delay, age, and stroke severity on the effects of intravenous thrombolysis with alteplase for acute ischaemic stroke: a meta-analysis of individual patient data from randomised trials. Lancet. 2014;384(9958):1929–1935.

24. Chen CH, Tang SC, Chen YW, et al. Effectiveness of Standard-Dose vs. Low-Dose Alteplase for Acute Ischemic Stroke Within 3-4.5 h. Frontiers in neurology. 2022;13:763963.

25. Wang D, Pocock S. A win ratio approach to comparing continuous non-normal outcomes in clinical trials. Pharmaceutical statistics. 2016;15(3):238–245.

26. Kaur J, Tuor UI, Zhao Z, Barber PA. Quantitative MRI reveals the elderly ischemic brain is susceptible to increased early blood-brain barrier permeability following tissue plasminogen activator related to claudin 5 and occludin disassembly. Journal of cerebral blood flow and metabolism : official journal of the International Society of Cerebral Blood Flow and Metabolism. 2011;31(9):1874–1885.

27. Charidimou A, Turc G, Oppenheim C, et al. Microbleeds, Cerebral Hemorrhage, and Functional Outcome After Stroke Thrombolysis. Stroke. 2017;48(8):2084–2090.

28. Chen J, Zeng Z, Fang Z, Ma F, Lv M, Zhang J. Risk factors for thrombolysis-related intracranial hemorrhage: a systematic review and meta-analysis. Thrombosis journal. 2023;21(1):27.

29. Keyt BA, Paoni NF, Refino CJ, et al. A faster-acting and more potent form of tissue plasminogen activator. Proceedings of the National Academy of Sciences of the United States of America. 1994;91(9):3670–3674.

30. Waseem MH, Abideen ZU, Khan MH, et al. Comparative Efficacy and Safety of Different Tenecteplase Doses With Alteplase in Acute Ischemic Stroke: A Systematic Review With Pairwise and Network Meta-Analysis to Determine the Optimal Dose. Brain and behavior. 2025;15(8):e70756.

